# A Framework for Falsifiable Explanations of Machine Learning Models with an Application in Computational Pathology

**DOI:** 10.1101/2021.11.01.21265738

**Authors:** David Schuhmacher, Stephanie Schörner, Claus Küpper, Frederik Großerueschkamp, Carlo Sternemann, Celine Lugnier, Anna-Lena Kraeft, Hendrik Jütte, Andrea Tannapfel, Anke Reinacher-Schick, Klaus Gerwert, Axel Mosig

**Affiliations:** Ruhr-University Bochum, Center for Protein Diagnostics Gesundheitscampus 4, 44801 Bochum; Ruhr-University Bochum, Faculty of Biology and Biotechnology, Bioinformatics Group Universitätsstr. 150, 44801 Bochum; Ruhr-University Bochum, Faculty of Biology and Biotechnology, Department of Biophysics Universitätsstr. 150, 44801 Bochum; Institute of Pathology, Ruhr-University Bochum, Bochum, Germany; Department of Hematology, Oncology and Palliative Care St. Josef Hospital, Ruhr-University Bochum, Bochum, Germany

## Abstract

In recent years, deep learning has been the key driver of breakthrough developments in computational pathology and other image based approaches that support medical diagnosis and treatment. The underlying neural networks as inherent black boxes lack transparency, and are often accompanied by approaches to explain their output. However, formally defining explainability has been a notorious unsolved riddle. Here, we introduce a hypothesis-based framework for falsifiable explanations of machine learning models. A falsifiable explanation is a hypothesis that connects an intermediate space induced by the model with the sample from which the data originate. We instantiate this framework in a computational pathology setting using label-free infrared microscopy. The intermediate space is an activation map, which is trained with an inductive bias to localize tumor. An explanation is constituted by hypothesizing that activation corresponds to tumor and associated structures, which we validate by histological staining as an independent secondary experiment.

## 1 Introduction

Classifications obtained from machine learning models in medical applications are in general black-box decisions, and interpreting their outcome in is often considered key for making machine learning based diagnoses or treatment decisions transparent and trustworthy [Abels et al., 2019, Kelly et al., 2019, Holzinger et al., 2019, Rudin, 2019]. The power of deep learning based classification has become particularly evident in image-based diagnostic settings, most notably in radiology and pathology [Litjens et al., 2017, van der Laak et al., 2021], where the need for interpretation has commonly been addressed by approaches such as linear relevance propagation (LRP) [Bach et al., 2015, Lapuschkin et al., 2019], saliency maps [Simonyan et al., 2013], Grad-CAM [Selvaraju et al., 2017] or class activation maps [Zhou et al., 2016]. These methods operate in a post-hoc manner in the sense that they operate on the neural network after it has been trained. Using the trained network along with an input image, they yield segmentations or heatmap representations [Zech et al., 2018, Hägele et al., 2020, Ding et al., 2019, Korbar et al., 2017, Bai et al., 2020, Hosny et al., 2018] that localize relevant patterns and can be assessed by domain experts. However, while unveiling an interpretable output, existing approaches lack a clear definition what constitutes a valid interpretation or explanation.

Ever since Lipton diagnosed the notion of interpretability to be ill-defined [Lipton, 2018], several attempts have been made to define interpretability and explainability in the context of machine learning models, as surveyed in Roscher et al. [2020], Guidotti et al. [2018], Holzinger et al. [2019] and recently in Samek et al. [2021]. However, it has been stated explicitly that no consensus definition has emerged, neither in a general sense [Guidotti et al., 2018] nor specific to biomedical image analysis [van der Laak et al., 2021]. Montavon et al. [2018] define an interpretation as *a mapping of an abstract concept into a domain that the human can make sense of*, while an explanation is *the collection of features of the interpretable domain, that have contributed for a given example to produce a decision*. The lack of a clear definition what an interpretation is has also been noted by Murdoch et al. [2019], who embed an interpretation in a so-called data-science life cycle and introduce the *predictive, descriptive, relevant* framework as a basis for interpretable machine learning. Despite these attempts towards definitions, apparently none has emerged as a consensus.

The challenge to define interpretability and explainability in the context of experimental research stems from the seemingly dichotomous approaches behind experimental research and machine learning. Experimental research, on the one hand, is hypothesis-centric by following hypothetico-deductivism, where predictions derived from a general and falsifiable hypothesis are applied to specific cases that either corroborate or falsify the hypothesis. Machine learning, on the other hand, derives general models from specific training examples and thus follows inductive [Wolpert, 1996] (or sometimes transductive [Vapnik, 2006]) reasoning. In machine learning, human expert-centric studies are common and legitimate practice in the sense that models are trained inductively on ground truth labels obtained from a human expert or a panel of human experts [Bulten et al., 2020, Abels et al., 2019, Amgad et al., 2019, Sirinukunwattana et al., 2017]. In the context of deductive experimental research, however, it appears reasonable if not inevitable to define explainability in a hypothesis-centric manner rather than an expert-centric manner. The early and remarkable work by Michalski [1983] on concept learning deserves credit for assigning the central role to hypotheses and explicitly addressing the problem of induction. Yet, Michalski’s approach of learning structural descriptions from examples lacks an explicit role of validating hypotheses in a deductive manner.

Here, we introduce an abstract and generic framework for outcome interpretation that is built upon a falsifiable hypotheses as the central constituent. The framework involves a machine learning model whose output yields, besides a classification outcome, an interpretable domain which is explicitly inferred during training of the model. The underlying hypothesis is formulated as a connection between the interpretable domain and the sample from which input data to the classifier have been obtained. Since the hypothesis refers to the sample rather than the input data, specific predictions about the sample can be derived from the hypothesis, which can thus be validated, i.e., corroborated or falsified, by additional experiments.

We demonstrate the application of our framework in a computational pathology setting dealing with colorectal tissue samples. Specifically, we introduce a weakly supervised deep convolutional neural network approach for classifying cancer and localizing tumor in microscopic images of tissue thin sections. We primarily work on hyperspectral infrared (IR) microscopic images which at each pixel position provide a location-specific biochemical fingerprint of the sample with a spatial resolution of around 5*µ*m [Kallenbach-Thieltges et al., 2013, Raulf et al., 2020]. Our *comparative segmentation network* approach (CompSegNet) is trained on sample labels only, but during training infers a segmentation of tumor which facilitates a hypothesis-based interpretation of classification outcome.

## 2 Theory and Calculations

### 2.1 A formal definition of model explanation

The computational approaches introduced here emerge from a formal definition of model interpretation in machine learning that is illustrated in Figure 1. We assume a given machine learning model *f* that classifies input *x* ∈ *X* derived from a specific sample as label *y* = *f* (*x*). In this setting, interpretation requires an extension of the machine learning model to infer additional interpretable output, so that *f* (*x*) = (*y, h*) with *h* ∈ *I*. Here, *I* is an *interpretable* space that is inductively inferred while (or subsequent to) training *f*. Formally, the extension is thus a mapping *f* : *X* →*Y* ×*I*, where *X* is the space of measured data, *Y* the space of supervised training labels, and *I* is what will henceforth be referred to as *interpretable space*, or I-space for short. This allows us to define an *explanation* of *f* (*x*) = (*y, h*) as a falsifiable hypothesis that establishes a connection between *h* ∈ *I* and the sample from which data *x* have been obtained. We will refer to hypotheses satisfying this definition as *falsifiable explanations* of the underlying machine learning model. The definition can be restricted further by requiring the classification *y* to be obtained purely on the grounds of *h* ∈ *I*. Formally, this means that classification is performed in two steps, where first *h* = *f* (*x*) and subsequently *y* = *g*(*h*) is computed. In this case, we refer to the space *I* as an *intermediary I-space*, and correspondingly define a hypothesis connecting *h* with the original sample as a *falsifiable intermediary explanation* of the classifier *g* ○ *f*. Intermediary explanations are obviously much stronger, since the classification can be derived from the interpretable intermediate variable, while the less restricted general falsifiable explanations allow *h* to be computed fully independent from *y*. Also, post-hoc interpretations [Bach et al., 2015, Simonyan et al., 2013, Zhou et al., 2016, Selvaraju et al., 2017] can be phrased in this framework. If the machine learning model is described by *M* in the space of all trainable models, its output can be denoted by *f*_*M*_ (*x*). The post-hoc interpretable space for input *x* is then computed through a function *h* = *g*(*M, x*), and defining *f* (*x*) := (*f*_*M*_ (*x*), *g*(*M, x*)) matches the framework displayed in Figure 1.

**Figure 1:**
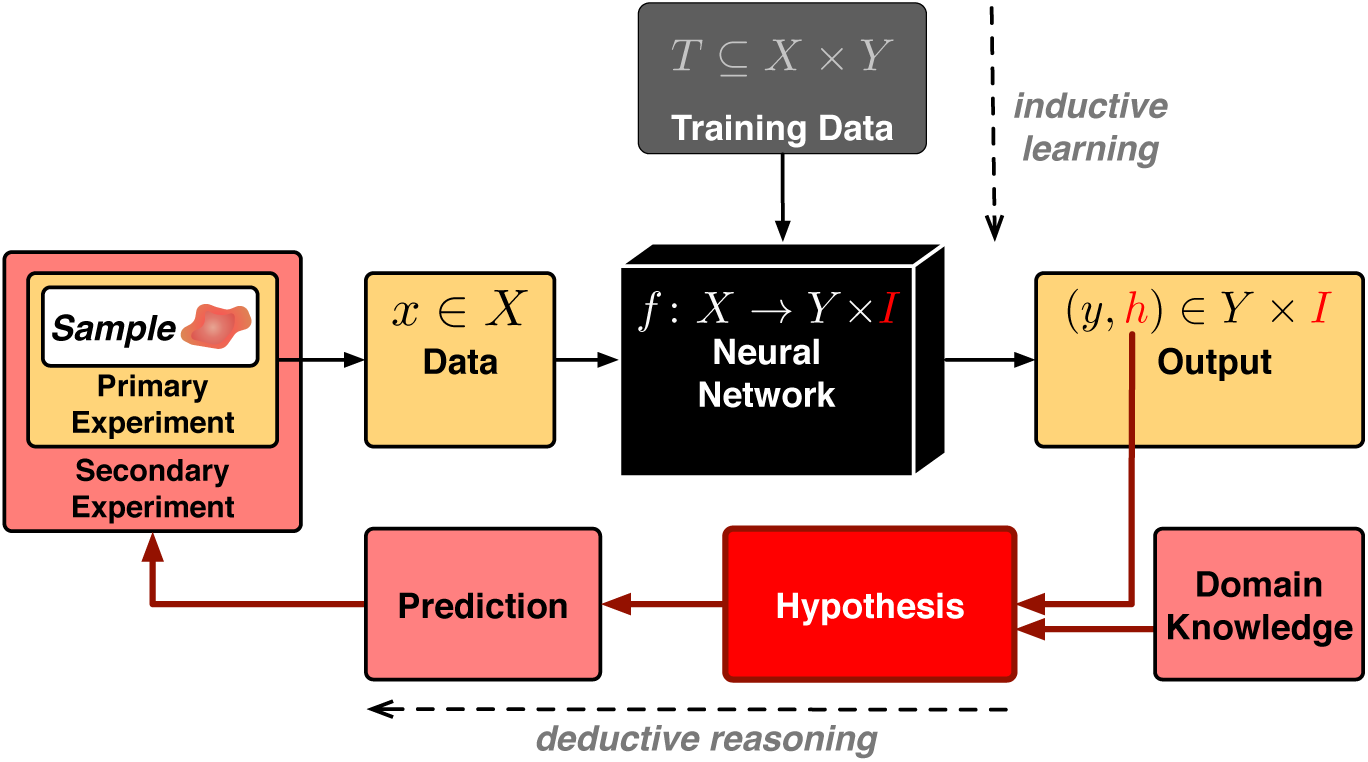
Framework for a formal definition of machine learning model explanation. Conventionally, supervised training of a machine learning model *f* based on measurements obtained from data space *X* equipped with labels *Y* is an inductive step. Interpretation is facilitated by extending *f* to infer an additional output, so that *f* (*x*) = (*y, h*), where *y* ∈ *Y* is a label and *h* ∈ *I* is additionally inferred output in an interpretable space *I*. A *falsifiable explanation* then is a hypothesis that connects *h* with the sample from which *x* originates.

There are crucial differences between falsifiable explanations and earlier attempts [Montavon et al., 2018, Murdoch et al., 2019] to define explainability and interpretability. First and foremost, our approach refers back to the physical reality of the sample from which the data originate rather than just referring back to the data *x* ∈ *X*. Second, we explicitly require an explanation to be a falsifiable hypothesis. Earlier definitions of explainability commonly involve understandability by a human expert. Our definition shifts understandability towards the hypothesis, which is implicitly assumed to be understandable by a human expert. Compared to [Michalski, 1983], the hypothesis is phrased ex-silico and outside the formalism of statistical learning, but within the rigor of hypothetico-deductive reasoning.

### 2.2 A neural network approach facilitating falsifiable explanation

We present the *comparative segmentation network* (CompSegNet) as a weakly supervised neural network that yields an I-space through an activation map which is trained on image labels only. A preliminary version of the CompSegNet has been described earlier [Schuhmacher et al., 2020]. Here, we extend the CompSegNet to a falsifiable explainable model in the context of whole-slide infrared microscopic images of histopathological tissue samples. In particular, we showcase how the CompSegNet yields an I-space and facilitates a falsifiable intermediary explanation of a classifier that distinguishes cancer samples from cancer-free samples.

As illustrated in Figure 2, the CompSegNet is a U-Net [Ronneberger et al., 2015] extended by a pooling neuron which accumulates activation from the output layer of the U-Net. Applied to distinguishing cancerous samples from cancer-free samples, the loss function aims to maximize pooled activation in images labeled as cancerous, while minimizing pooled activation in images labeled as cancer-free. This loss function invokes an inductive bias for the U-Net to localize structures that are present in cancer samples only. In other words, we may *hypothesize* that the CompSegNet localizes tumor and associated structures. It must be emphasized that the I-space is the outcome of an inductive process and thus formally decoupled from epistemic concepts such as tumor. The concept of tumor is unmistakably characterizable on a molecular basis. The hypothesis connects the inductively obtained I-space with epistemic concepts from molecular reality, thus resolving the epistemological opacity [Durán and Jongsma, 2021] of the model.

**Figure 2:**
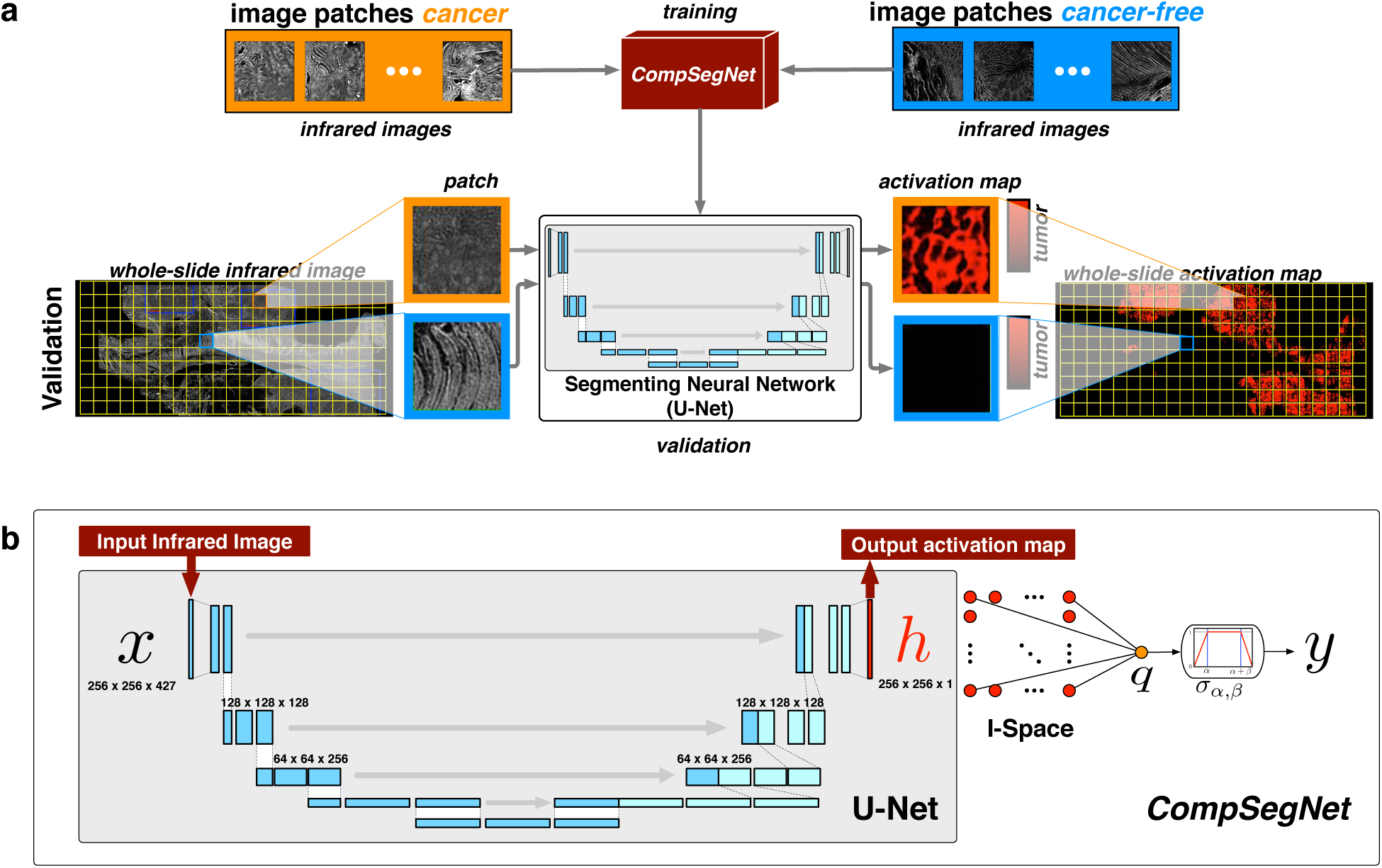
Schematic overview of the Comparative Segmentation Network (CompSegNet). **a** The network is trained on sample labels only, but yields a U-Net [Ronneberger et al., 2015] whose output layer as an activation map facilitates segmentation. **b** The activation map is inferred with the help of a pooling neuron whose activation is maximized for cancer samples and minimized for control samples during training.

The hypothesis that activation in I-space corresponds to tumor, henceforth referred to as *tumor-activation hypothesis* for short, is indeed falsifiable. In the given setting, images have been obtained from label-free infrared microscopy, which is particularly amenable to further experimental characterization since the sample is unaltered after infrared microscopic imaging. Specifically, as a secondary validation of our hypothesis, we performed conventional hematoxylin and eosin (H&E) staining, where tumor regions can be clearly identified and the hypothesis can thus be validated.

### 2.3 Network Architecture

We formally consider a microscopic image *x* involving *d* optical channels as a mapping *x* : Ω ×[1 : *d*] → ℝ in a coordinate system Ω with *N* × *N* pixels. We consider conventional light microscopic H&E stained images involving *d* = 3 channels and hyperspectral infrared images where each pixel is represented by a spectrum involving *d* = 427 wavenumbers. In a complete data set Χ = {(*x*_1_, *y*_1_), …, (*x*_*n*_, *y*_*n*_) }, each image *x*_*i*_ is associated with a sample-level label *y*_*i*_, with *y*_*i*_ = 1 indicating a diseased sample, and *y*_*i*_ = 0 indicating a healthy control sample. As a notational convention, we denote the optical spectrum at position *j* ∈ Ω by *x*_*i*_(*j*). The goal of training our network is to infer → an activation map *h*_*i*_ : Ω → [0, 1] that yields an I-space by localizing disease patterns in the diseased samples: If *x*_*i*_ is a cancer sample indicated by *y*_*i*_ = 1, the output map *h*_*i*_(*j*) should approach 1 precisely for those positions *j* where tumor or tumor-associated tissue is present, whereas at all tumor-free positions *j* in *x*_*i*_, *x*_*i*_(*j*) should approach 0. In particular, given a healthy control image *x*_*i*_, *all* positions should receive an activation close to 0 in their activation map *h*_*i*_.

Our network is essentially an extended U-Net architecture [Ronneberger et al., 2015] that receives as input an image *x*_*i*_ and produces the activation map *h*_*i*_ as an I-space variable from which the output of the network function *f* (*x*_*i*_) is computed through a pooling neuron. Beside a modified transfer function in the activation map and the extended optical dimensionality of the input, we employed the widely established topology of the U-Net. For the conventional H&E data, we use the U-net topology from Ronneberger et al. [2015] with an input dimensionality of 224 × 224 × 3. In the experiments with the hyperspectral image data, the input size was adapted to 256 × 256 with 427 optical channels at each pixel, while the remaining layers of the network were unchanged.

The topology, parameter space and loss function of the network are expliciteply tailored to accomodate modeling assumptions that adjust the inductive bias of the training process. In particular, to accomodate the nature of histopathological imaging data, positions in the sample that are not covered by sample need to be disregarded in the loss function. To this end, we introduce a binary background mask *M*_*i*_. We indicate *M*_*i*_(*j*) = 1 if pixel *j* in image *i* is covered by sample, and *M*_*i*_(*j*) = 0 otherwise. For both infrared microscopic images and H&E stained images, *M*_*i*_ can be computed in a straightforward manner from *x*_*i*_. For the infrared microscopic images, a threshold is calculated based on the histogram of the absorbance at 1656 *cm*^−1^ over the whole image using Otsu’s binarization approach with the use of a gaussian filter of size 5. For H&E images, positions not covered by sample appear bright white and can be similarly separated by a intensity threshold.

For the interpretation of network output, it is essential that the activation map *h*_*i*_(*j*) at each position *j* is bounded within the interval [0, 1]. To achieve this, a sigmoid activation function is used for each neuron in the neural network layer representing *h*. The activations of the neurons that constitute the activation map *h*_*i*_ are each weighted by 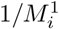, with 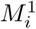 representing the number of foreground pixels in the background mask *M*_*i*_ and then accumulated in a pooling neuron *q*; positions masked as background are associated with zero activation. A key modeling ingredient is the transfer function *σ*_*α,β*_ for the pooling neuron *q*. The transfer function is used to steer the network towards identifying a certain minimum and maximum fraction of tumor in input images labeled as cancer. At the same time, the transfer function needs to deincentivize the overdetection of cancer. Otherwise, without limiting overdetection, the network will escape towards the trivial solution and label each cancer sample completely as tumor. We translate these assumptions into a piecewise linear transfer function

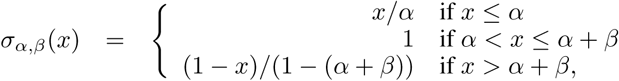

which puts a maximum incentive for the network to identify a fraction of tumor that lies in between *α* and *α* + *β* where *α* and *β* are two parameters with an explicit modeling role. In our present networks, we used *α* = 0.05 and *β* = 0.9 for the infrared image based CompSegNet and *α* = *β* = 0.4 for the H&E image based CompSegNet.

Through the pooling neuron *q* a segmenting network becomes a binary classifier, whose classification performance is based on the segmentation resulting from the activation layer. Thus, in case of a successful classification by the pooling layer, the corresponding pixel-precise segmentation can be extracted.

The contribution *L*^*i*^ of image *x*_*i*_ to the loss function used is composed of two terms

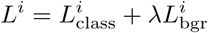

coupled by a scaling parameter *λ* that was constantly set to 1 throughout all computations presented here. The first part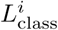 is a binary cross entropy with a class weighting. The class weights *w*_−_ and *w*_+_ were determined by the relative prevalence of positive and negative samples in the dataset. For an input image *x*_*i*_, we denote its given disease label as *y*_*i*_ and the output of the network after the pooling neuron as *f* (*x*_*i*_). Then, we write the class-loss as

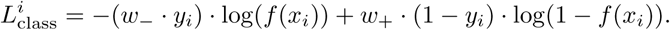

Activation of pixels that are either masked out as background through the mask *M*_*i*_ or belong to a negative cancer-free dataset is penalized by the term 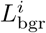 through

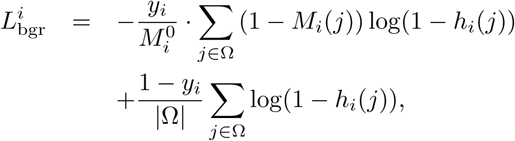

where 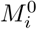 denotes the number of background pixels in mask *M*_*i*_. All networks were trained using a dropout rate of 20% in the hidden layers along with batch normalization and RMSprop as optimizer.

### 2.4 Activation Map Binarization

In order to binarize activation maps for computing Dice scores, we follow an approach that involves thresholds at two different levels. The primary threshold *θ* constitutes the actual activation threshold to binarize the activation map. A secondary threshold *ϱ* is a tumor-fraction threshold such that a sample is classified as cancer whenever the ratio of tumor pixels exceeds *ϱ* after binarization using *θ*. Now, each combination (*θ, ϱ*) turns the activation layer of a CompSegNet-trained U-Net into a classifier whose performance can be measured on the validation set by computing the F1_*θ,ϱ*_-score. Since both *θ* and *ϱ* are [0, 1]-valued, we can determine (*θ*_opt_, *ϱ*_opt_) = arg max_*θ,ϱ*_ *F* 1_*θ,ϱ*_ using exhaustive search with a step size of 0.02 for *θ* and a step size of 0.01 for *ϱ*.

## 3 Material and Methods

### 3.1 Infrared Imaging

Infrared imaging is a label-free method for obtaining robust and observer-independent classifications of tissue thin sections on microscopic spectral images. Based on the molecular fingerprint in the spectral domain, supervised learning algorithms can be trained to identify disease-associated regions.It has been shown that this method allows adiffer-entiation of tissue types with focus on cancer tissue sections [Kuepper et al., 2018]. It also allows the biochemical differentiation of tumor tissue in terms of cancer subtyping and grading [Großerueschkamp et al., 2015, Kuepper et al., 2016].The original setup was based on Fourier transform infrared spectroscopy (FTIR), embedded in a microscope setup. Due to long acquisition times a transition to Quantum Cascade Laser (QCL) based imaging approaches has been made which reduced the measurement time from 90 hours for the FTIR setup to approximately one hour foran average colon tissue thin section of 1.4 cm × 2 cm and yields high-quality spectral data [Kuepper et al., 2018]. As a QCL based platform, the Spero QT IR microscope (Daylight Solution, San Diego, CA, USA) was utilized.The standard configuration covers the wavelength from 1800 *cm*^−1^;1to 950 *cm*^−1^ with a spectral resolution of 2 *cm*^−1^. The installed 4 × 0.3 NA objective was used with a coverage of 2 × 2 *mm* Field of View (FOV). The uncooled microbolometer focal plane array (FPA) detector consists of 480 × 480 pixels with a pixel size of 4.25 × 4.25 *µm*. During the measurement the sample chamber is constantly flooded with dry air.

### 3.2 Patients and datasets: IR Imaging

The primary basis for our infrared microscopy based study is a collective of 100 samples of patients with colorectal carcinoma (UICC Stage II & III, older than 18 years) and 100 samples of tumour-free tissue samples, both randomly selected from the multicenter registry study ColoPredict Plus 2.0. The nation-wide study involves 188 clinical trial centers and collects samples retro- and prospectively. The 100 samples per group (tumor / tumor-free) were divided into 50% training data, 25% validation data and 25% independent test data, as shown in Table 1. The cohorts were balanced by gender, UICC Stage and localization with the whole cohort showing a tendency for a higher proportion of female patients, Stage II and a right-sided localization. For some samples, the data set was extended by measurements on a second microscope of same type to include device variance in the dataset, so that in total, the study is based on 200 samples and 488 IR measurements of 190 patients. For a detailed description of the cohort refer to Kallenbach-Thieltges et al. [Kallenbach-Thieltges et al., 2020]. The subdivision into training, validation and test group was performed strictly at the patient level.

**Table 1:** Patient characteristics for the infrared imaging dataset from the ColoPredict Plus 2.0 multicenter cohort. The total numbers are provided for the number of samples (N(s)), the number of measurements (N(m)) and the number of patches (N(patch)). The data provided for the characteristics are the numbers and the percentages corresponding to N(s). For the tumor-free samples only data about gender and age can be provided.

|  | Training Cohort |  | Validation Cohort |  | Test Cohort |  |
| --- | --- | --- | --- | --- | --- | --- |
|  | *Tumor* | *Tumor-free* | *Tumor* | *Tumor-free* | *Tumor* | *Tumor-free* |
| **Total Number** |  |  |  |  |  |  |
| *N(s)* | 50 | 50 | 25 | 25 | 25 | 25 |
| *N(m)* | 137 | 106 | 73 | 54 | 59 | 59 |
| *N(RoI)* | 650 | 645 | 318 | 320 | 240 | 364 |
| **Gender** |  |  |  |  |  |  |
| *female* | 32 (64%) | 27 (54%) | 15 (60%) | 8 (32%) | 14 (56%) | 13 (52%) |
| *male* | 18 (36%) | 23 (46%) | 10 (40%) | 17 (68%) | 11 (44%) | 12 (48%) |
| **Age in years** |  |  |  |  |  |  |
| mean ± SD | 77±11.9 | 68±10.6 | 76±13.2 | 72±9.5 | 73±10.4 | 71±10.2 |
| **UICC Stage** |  |  |  |  |  |  |
| II | 30 (60%) | n.a. | 15 (60%) | n.a. | 14 (56%) | n.a. |
| III | 20 (40%) | n.a. | 10 (40%) | n.a. | 11 (44%) | n.a. |
| **Localization** |  |  |  |  |  |  |
| *left* | 18 (36%) | n.a. | 7 (28%) | n.a. | 11 (44%) | n.a. |
| *right* | 32 (64%) | n.a. | 18 (72%) | n.a. | 14 (56%) | n.a. |

The samples were obtained during surgery, locally formalin-fixed and paraffin-embedded and subsequently treated according to the standard procedure at the Institute of Pathology. Samples were cut into 7 *µ*m thick tissue thin sections following a previously established infrared microscopy workflow [Kuepper et al., 2018] described in Section 3.1. Subsequent to infrared imaging, the tissue thin sections were stained by H&E following standard procedures in pathology. Based on the H&E stained tissue sections, each measurement was annotated on a high-level scale by a pathologist, marking regions slightly larger than the input patch size of the CompSegNet (≈ 1.1 *mm*) that contain a minimum of 20% tumor-related tissue in the 100 tumor samples and checking the tumor-free samples. As a data augmentation strategy, patches for training were obtained by randomly cropping patches matching the CompSegNet input size of 256×256 pixels. For further augmentation, the patches are flipped randomly after cropping.

In order to validate the tumor-activation hypothesis associated with the trained CompSegNet, three tissue sections were H&E stained as secondary experiment. In the resulting images, tumor-associated regions were annotated by a pathologist, yielding a binary mask with one class representing the tumor-associated tissue regions and the other class representing all tumor-free regions which includes tissue components such as crypts, lymphoid follicles, connective tissue, blood vessels and nerve tissue as well as muscle. The annotation of tumor-associated regions was performed in a broad sense, including regions which may not contain tumor cells, but do not occur in tumor-free tissue sections, which is generally referred to as stroma. This includes, for example, the extracellular matrix, immune cells, infiltrated connective tissue and basement membranes.

### 3.3 Dataset: H&E Images

For further validation of the CompSegNet approach, the H&E data set from Kather et al. [2019a], Kather et al. was utilized for training and validation as a well-established data set in computational pathology. It provides tissue samples from the National Center for Tumor diseases (NCT, Heidelberg, Germany) tissue bank and the University Medical Center Mannheim (UMM, Heidelberg University, Mannheim, Germany). The larger dataset provides 100,000 non-overlapping image patches from 86 H&E stained slides scanned by a conventional whole-slide scanning microscope and is referred to as *NCT-CRC-HE-100K*. Each image patch is 224 × 224 pixels in size at 0.5 *µ*m per pixel and is assigned one out of nine different classes, with two classes associated with colorectal cancer (cancer-associated stroma and colorectal adenocarcinoma epithelium). The smaller dataset is referred to as *CRC-VAL-HE-7K* and consists of 7180 image patches, with the same size and resolution as the large data set.

## 4 Results

### 4.1 Training performance

For the application of the CompSegNet to infrared data, the number of input channels was extended to 427 optical channels. The expected tumor fraction thresholds, as described in chapter 2.3, were set to *α* = .05 and *β* = .9, matching the expected rate of tumor content within an image patch and thus controls the inductive bias of the network based on modeling assumptions. The dropout rate was set to 20% along with a learning rate of 5·10^−4^. The RMSprop parameters were set to *ρ* = 10^−6^ with a momentum of 0. For the learning rate, a scheduler was implemented with a decay of 0.9 every 30 epochs along with a batch size of 4.

Figure 3 a illustrates the loss for the training and validation dataset over the 250 epochs of training. The first 6 epochs show a slowly dropping plateau phase. This is a consistent behaviour of the CompSegNet and is explicitly guided by a background recognition term in the loss function. The length of the plateau phase is inversely proportional to the learning rate. In addition to the loss, a visual illustration has been added for five epochs over the course of training that shows an overview of the predictions of the validation patches in the I-space, where the evolution of the inferred I-space can be traced over training time. The evolution of the learned I-space can also be traced visually in the Supplementary Video. The final epoch (epoch 244) has been selected with regard to the area under receiver operating characteristic (AUC) values for the receiver operating characteristic (ROC), the F1 score, sensitivity, specificity and accuracy on the validation dataset. Validation on the independent test set exhibits similar performance as on the validation set as indicated in Figure 3, indicating a stable generalization performance.

**Figure 3:**
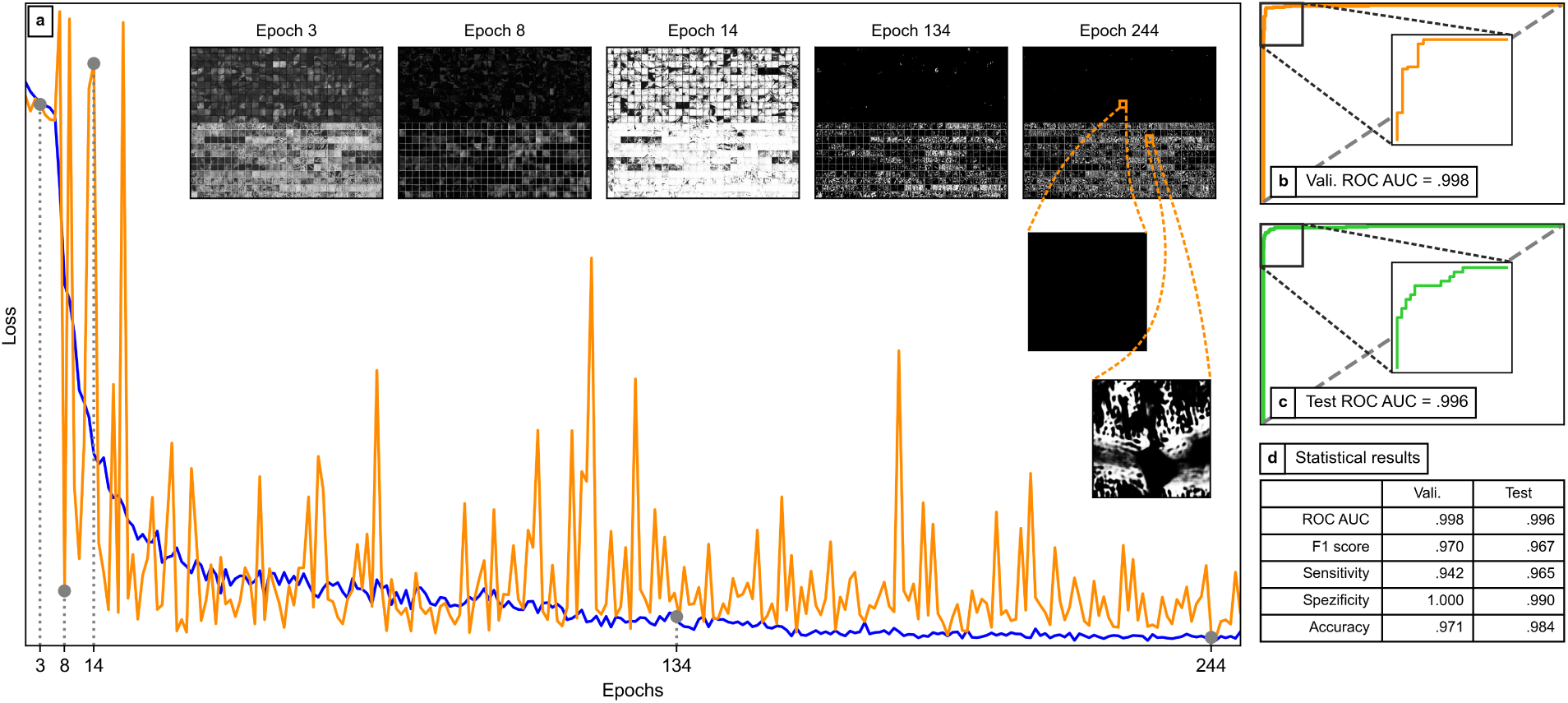
Training curve, evolution in I-space and ROC curves of a CompSegNet trained on infrared images of colorectal histopathology tissue sections with patch size of 256 × 256 pixels. **a** Training progress for IR images with overview-images: The blue and orange curve show the training and validation loss over 250 epochs. Five epochs were chosen as examples to visually illustrate the course of training in the I-space for the validation data. The upper half of the five overview images displays activation within the tumor-free patches and the lower half within the tumor patches. **b-c** ROC curves and AUC values for validation (N(patch)=638) and test dataset (N(patch)=604) for the selected model from epoch 244. **d** The table shows all statistic parameters that were used for selecting the model.

### 4.2 Validating the tumor-activation hypothesis

In order to validate the tumor-activation hypothesis, we applied the trained CompSegNet to a set of three whole-slide images from the independent test set.The prediction for the whole-slides was performed by moving a 256 × 256 pixel window over each measurement with an overlap of 64 pixels while assigning maximum activation value in the overlapping areas. The resulting whole-slide activation map was binarized by a global activation threshold of .628 as described in chapter 2.4. Agreement between the resulting binarized mask and tumor annotations was measured using Dice scores after transferring the H&E image and its annotation into the same coordinate system as the infrared image using the registration approach from Trukhan et al. [2020]. Figure 4 a-i illustrates the results on the whole sample for three thin tissue RoI from the test cohort. The Dice scores consistently exceed .5 indicating a good agreement between tumor-associated regions and the inferred I-space. The I-space is consistently a subset of the annotated regions. This may at least partially be explained by the inherent practical limitations of annotating the tumor-associated regions, which is complicated by the reduced H&E image quality that is inherent to the PET microscopy slides used for infrared microscopy. For the purpose of linking back the I-space to the sample, two subregions from Figure 4 f were selected that show examples of both agreement and disagreement between I-space activation and annotation. By looking at the spectral property of the underlying tissue as shown in Figure 4 f (iii), a molecular interpretation becomes possible that closes the gap between I-space, hypothesis and the biochemical properties of the sample itself. For a high-level overview, two whole-slide H&E images are illustrated with the I-space activation in green in Figure 4 j and k, showing one cancer and one cancer-free sample, respectively.

**Figure 4:**
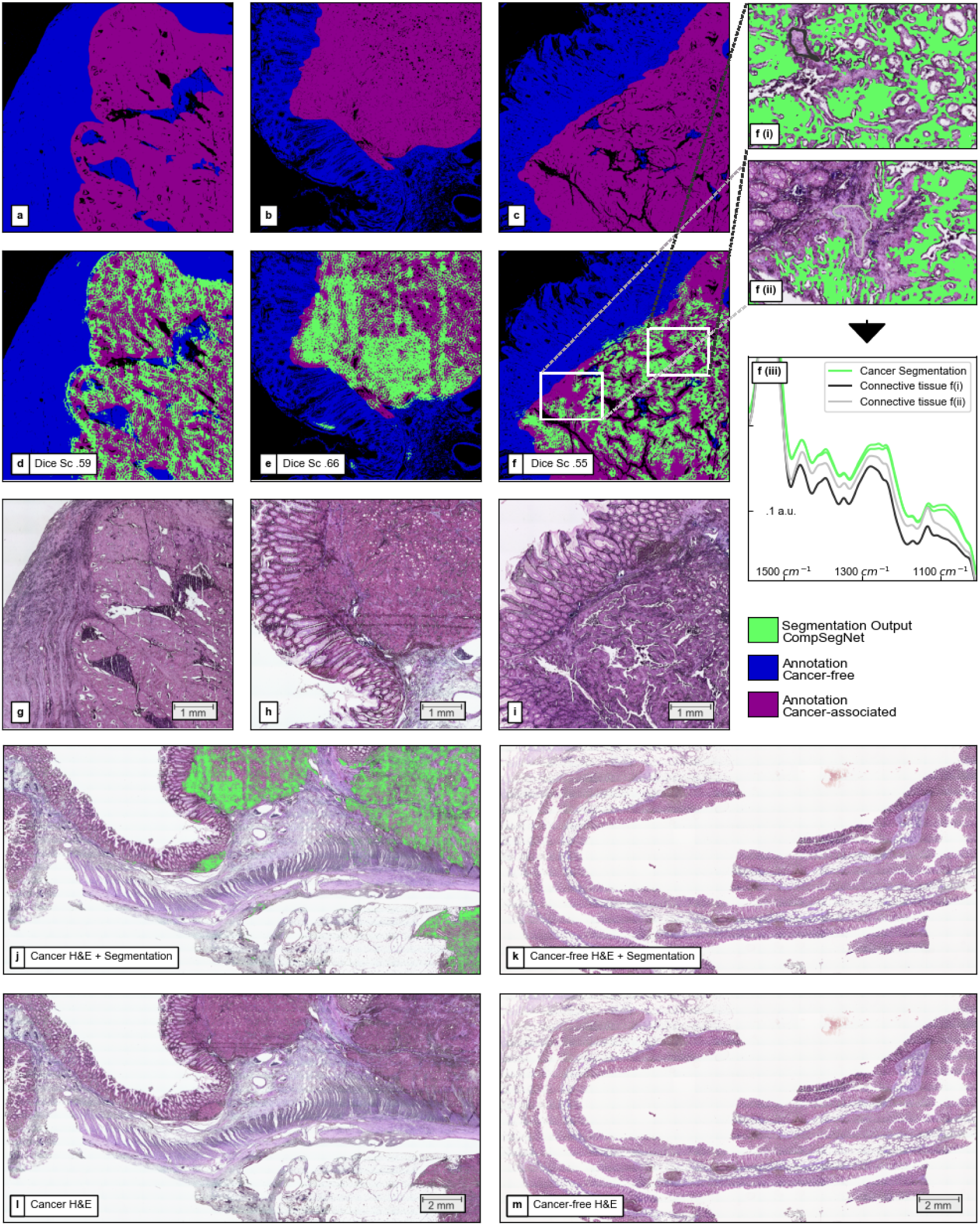
Results for whole-slide segmentation on infrared measured samples from the test cohort of the ColoPredict Plus 2.0 study. **a-c** Ground truth annotation of H&E images with regard to tumor-free regions and tumor-related regions. **d-f** show the annotation from a-c overlaid with the segmentation output of the CompSegNet in green. **f(i)** and **f(ii)** show a magnification of f for two selected regions in which tumor-associated tissue was annotated, but no activation of the network appeared. Infrared spectra are plotted for cancer segmentation areas and highlighted non-activated areas. For interpretation of annotation and segmentation, **g-h** shows the matching H&E stained sections for d-f. **j-m** illustrates two whole-slide H&E stained images, overlaid with the activation map (j: cancer-sample, k: cancer-free sample) and the corresponding H&E image without segmentation overlay (l: cancer-sample, m: cancer-free sample).

**Figure 5:**
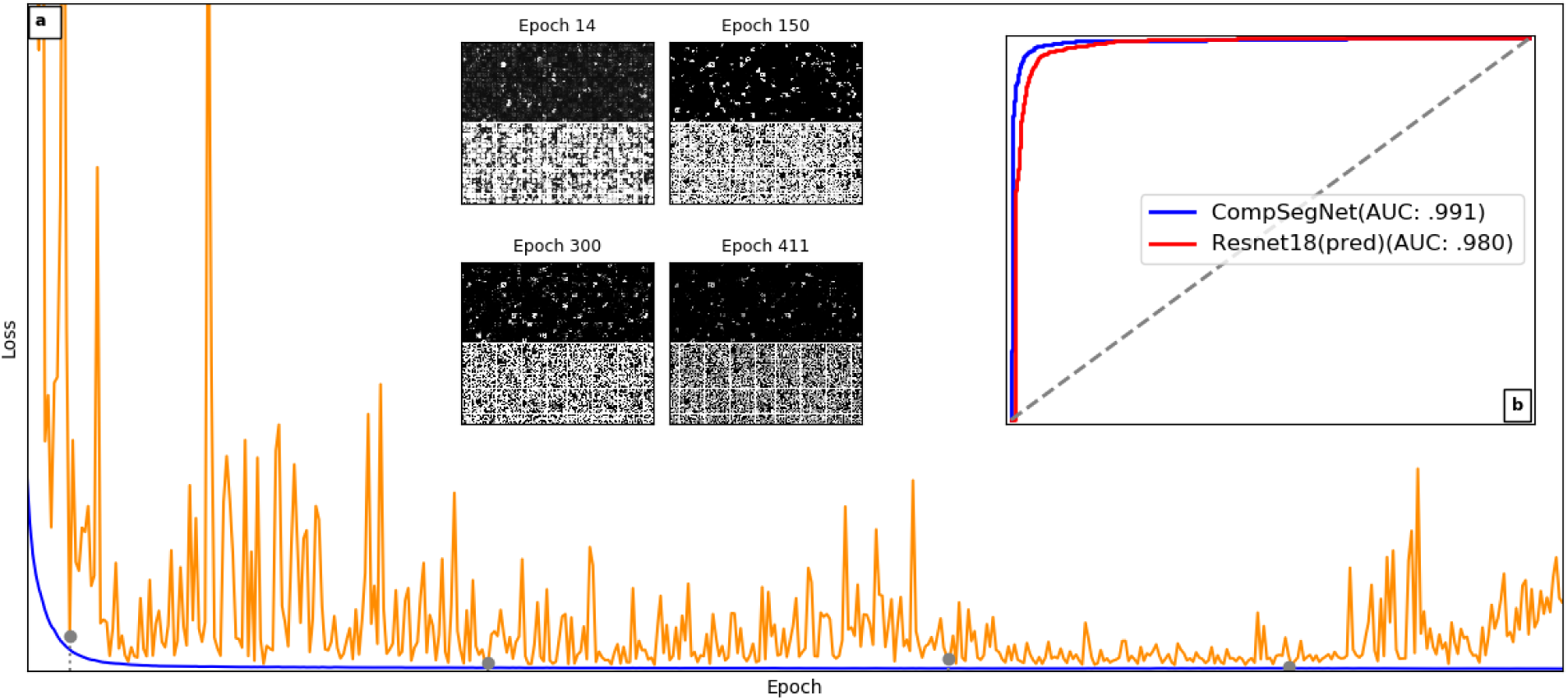
Training curve and ROC curve of a CompSegNet trained on H&E stained images of colorectral histopathology image data. **a** Loss during training are shown in blue and orange for training set and validation set, respectively. **b** The ROC curve was obtained based on the validation set from [Kather et al., 2019a], using all 1200 tumor patches against 1200 patches randomly sampled from the remaining 8 classes as a class-balanced validation set. A visual snapshot displaying the activation maps of a random subset of the validation data set is provided for four epochs. The ROC curve of the CompSegNet consistently exceeds the ROC curve of predictions through a ResNet-18.

### 4.3 Tumor segmentation in conventional histopathology image data

In order to further validate the CompSegNet approach, we trained a network to segment colorectal carcinoma related structures using the H&E data set from Kather et al. [2019b], as described in section3.3. As training data, the dataset provides 100,000 image patches from 86 H&E stained slides. Each image patch is 224 × 224 pixels in size and is assigned one out of nine different classes. In order to match the characteristics of the training data, we chose the expected tumor fraction parameters of the loss function as *α* = *β* = .4, corresponding to an expected tumor content of 40-80% in each patch annotated as tumor. The training data set is complemented by a validation dataset with about 7000 images with the same parameters. For independent testing, we further supplemented the data set with large subregions of three further whole-slide images from the ColoPredict Plus 2.0 registry study summarized in Table 1. In these three samples, tumor and associated regions were annotated as ground truth. All H&E stained images were normalized using the approach by Macenko et al. [2009]. Note that in the setting of applying the CompSegNet to H&E stained images, the H&E images themselves cannot be used as secondary experiment for validating the tumor activation hypothesis. Yet, in order to measure the agreement of inferred activation with tumor-associated tissue, we annotated tumor regions and measured the overlap with activation. Compared to the underdection observed in the infrared-based activation maps, the H&E based activation maps displayed in Figure 6 rather display over-detection in two out of three samples of tumor-related structure, yet with similar Dice scores. In the third sample (Figure 6 **c**), one part of the tumor is not recognized, potentially due to tumor structures not represented in the training data set.

**Figure 6:**
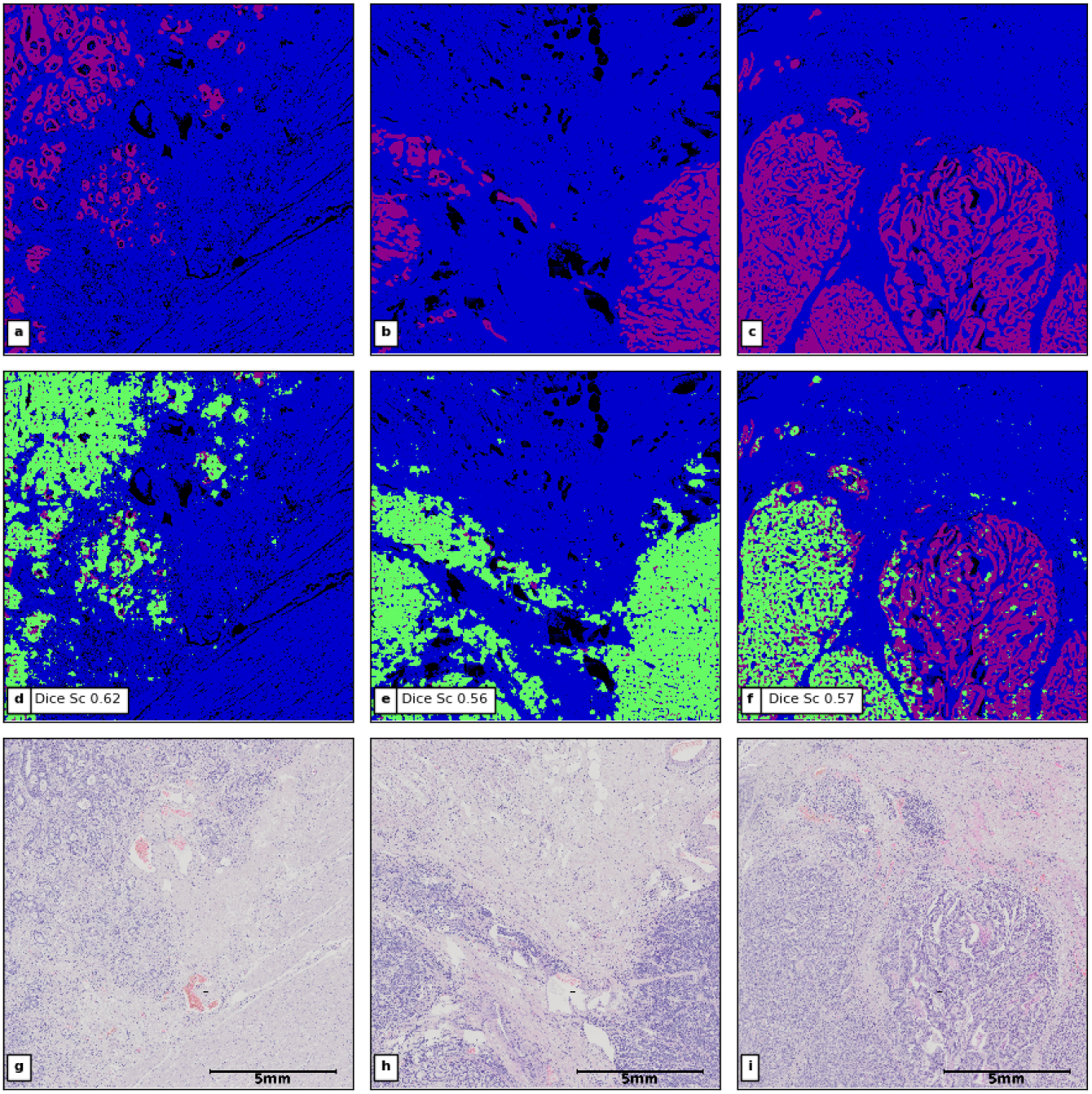
Segmentations of H&E stained images from the fully independent ColoPredict Plus 2.0 study test set. **a-c** Ground truth annotation of tumor and associated structures. Following Figure 4, annotated tumor-associated regions are shown in purple, other tissue in blue. **d-f** Ground truth overlaid with binarized segmentation map (green). **g-i** H&E staining images underlying the annotation and the activation map.

We compared the I-space based classification of the CompSegNet with LRP as a reference for post-hoc explanation methods [Hägele et al., 2020]. Conceptually, LRP exhibits a tendency towards satisfying the tumor-activation hypothesis because the relevance map is built on the invariant that accumulated relevance equals network output [Bach et al., 2015]. However, as a post-hoc approach, LRP lacks corresponding inductive bias during training. As detailed in Supplement A, we followed Kather et al. [2019b] and trained a ResNet-18 [He et al., 2016] on the same data as the CompSegNet, and subsequently performed LRP on the image patches from the validation set. The ResNet ouput was used to compute ROC curves on the validation set, which due to the relevance invariant is equivalent to a ROC curve over accumulated pixel relevances. The ResNet-18 exhibits slightly but consistently worse performance than the CompSegNet, indicating an effect of the inductive bias on optimizing I-space during training of the CompSegNet. The same trend is observed on the independent test set, where the LRP based segmentation achieves an average Dice score of .54 (Supplementary Figure 1), compared to an average of .58 obtained by the CompSegNet.

As to be expected from the use of sigmoidal as well as piecewise linear transfer functions, convergence of the CompSegNet (Figure 5 **a**) is less smooth compared to the ResNet-18 (Supplementary Figure 1). With the rapid forward-calculations of the U-Net underlying the CompSegNet, the evolution of the learned I-space can be traced visually during training without much computational effort. For the backward propagation underlying LRP, this is impractical due to the much higher demand in computational resources (Supplement).

## 5 Discussion and conclusions

We have introduced a framework for falsifiable explanations of machine learning models which establishes a hypothesis rather than an expert as the major constituent of an explanation. We introduce the concept of I-space as the link between hypothesis and machine learning model. As we demonstrate, a suitable I-space can be obtained by either an explicit inductive bias during training or by post-hoc methods. We showcase these concepts by establishing a tumor-activation hypothesis for deep neural networks that identify cancer in histopathological imaging data.

Several aspects need to be pointed out regarding our definition of a machine learning model explanation. First, it is crucial that the hypothesis refers back to the original ex-silico sample rather than the already observed in-silico data *x*. By referring back to the sample, the hypothesis can (and in general will) facilitate a prediction about the outcome of a secondary type of experiment performed on the same sample, rather than a prediction about the experiment that has been conducted to obtain *x*. Second, the definition introduces a new layer of validating a machine learning model. In addition to validating at the level of training labels, the explanation of the machine learning model can be validated by testing the stated hypothesis. In particular, the model may be invalidated by falsifying the hypothesis. Third, the definition unifies inductive machine learning with the hypotheto-deductivism of the scientific method, since the inductive task of supervised training is complemented by deductive reasoning: Pure supervised training merely interpolates between data points, and the inductive bias of a machine learning method determines how to interpolate. Our approach introduces I-space, which is also subject to interpolation, but the arbitrariness of interpolation is blocked by a falsifiable hypothesis and predictions derived from it. In this sense, phrasing inductive bias through I-space can be thought of and used as a modeling tool that controls how the model interpolates. The deductive ex-silico validation level legitimates to train a model that optimizes towards certain features of I-space representations. In other words, I-space can be used to constrain and control the inductive bias of training *f*.

Our definition links to existing concepts and approaches of explainable machine learning. In the taxonomy introduced in [Guidotti et al., 2018], interpretations matching our definition are outcome explanations since they explain the decision on a given object in contrast to model explanations which explain the internal logic of a machine learning model independent of a specific input. In general, any of the numerous and widely used model explanation approaches proposed in literature such as linear relevance propagation [Bach et al., 2015], saliency maps [Simonyan et al., 2013] or class activation maps [Zhou et al., 2016, Selvaraju et al., 2017] are potential sources of an I-space. These methods usually infer the computation of an input-specific interpretation in a post-hoc manner after a black box neural network has been trained. While such inferred interpretations may support a hypothesis that withstands experimental validation, post-hoc methods of output explanation do not exploit the potential to optimize I-space inference towards a specific hypothesis by modeling inductive bias during training. It is important to notice that our CompSegNet approach yields a readily trained U-Net, which can process input using fast parallel forward calculations. This provides a practical and crucial advantage over most post-hoc methods, which require backpropagation steps that are more difficult to parallelize.

Some existing machine learning models implicitly match parts of our definition of falsifiable explanations. For example, applying multiple instance learning in histopathology [Campanella et al., 2019, Lu et al., 2021, Ilse et al., 2018, Li et al., 2019] tiles whole-slide samples into a large number of patches, and the key task is to identify the few disease-relevant patches among this large number. The few positive instances claim to localize disease, which yields a basis for a falsifiable hypothesis. In other words, multiple instance learning in histopathology yields an I-space at the level of patches. In this view of multiple instance learning, our CompSegNet approach is a weakly supervised learning approach that provides an I-space in the same coordinate system as the input image. Both the CompSegNet approach and the variants of multiple instance learning used in computational pathology [Campanella et al., 2019] involve different types of target functions that are tailored towards localizing disease under certain side assumptions. This optimization under side assumptions indeed introduces the desired inductive bias while training the models. In more general terms, certain types of weakly supervised learning may provide a basis for falsifiable explanations.

It is noteworthy that experimental validation is an inherent part of falsifiable explanations. It facilitates formal inclusion throughout all phases of clinical studies. This appears relevant since on the one hand, explainable models are often considered an important corner stone for trustworthy machine learning in medical applications [Abels et al., 2019, Kelly et al., 2019], while on the other hand no formal definition of what an explanation is has been established to date. The question arises whether explanations that are not connected to falsifiable hypotheses can or should formally be part of clinical studies. The necessity of interpretation has recently been argued by Durán and Jongsma [2021], who identify epistemic opaqueness as the primary reason to distrust black box models. Since falsifiable explanations connect domain knowledge with machine learning models within the rigor of the scientific method, our proposed framework does establish epistemic transparency of the underlying machine learning model.

Neither the input space nor the I-space is limited in any way to image data, and the concept of falsifiable explanations carries beyond biomedical settings. While the input space is constituted by a given experimental setup, the only constraint on I-space is that it needs to connect to the hypothesis that constitutes the actual interpretation. It is important to realize that the hypothesis may and in general should involve further domain knowledge about the underlying sample. For example, the tumor-activation hypothesis involves knowledge about tumorgenesis. The same holds for the task of designing a secondary experiment. Examining the hypothesis as in our case by performing H&E staining on the sample involves background knowledge that tumor can be recognized by densely packed nuclei of certain morphology in particular regions of the sample. Other experiments may be imaginable for the same hypothesis, e.g. dissecting the areas with high activation using laser capture microdissection [Großerueschkamp et al., 2017, Selbach et al., 2021] and subsequently performing genome sequencing on the dissected sample material. A possible prediction derivable from the hypothesis would be to observe an accumulation of somatic mutations in the dissected areas compared to areas with no activation in I-space. In order to obtain robust molecular characterizations from laser microdissection, a highly precise localization of disease patterns is required. This is a major motivation behind our CompSegNet approach to localize disease at full input resolution, rather than at tile-resolution as obtained from multiple instance learning approaches.

When approaching more detailed and complex classification tasks, for instance the identification of cancer subtypes, it can be expected that all essential parts of a falsifiable explanation will get more complex and involved, ranging from the I-space to the hypothesis and the prediction. In some biomedical settings, for instance when dealing with non-invasive radiological imaging approaches, the role of the sample is taken by the patient under examination. Here, a hypothesis may predict the outcome of further diagnostic procedures performed on the patient.

Finally, our comparison between the weakly supervised CompSegNet approach and LRP as a post-hoc approach demonstrates that in general both post-hoc and inductive bias based approaches are feasible approaches to infer a relevant I-space. Although our validations are limited to two types of histopathologic imaging data, the CompSegNet approach naturally translates to a variety of other image modalities, in particular if controlling the inductive bias through the design of dedicated loss functions is considered as a modeling step. Conceptually, the CompSegNet approach aims to overcome post-hoc machine learning models and takes a step towards models that are more, although certainly far from fully, intrinsically interpretable. Combined with the concept of falsifiable explanations, the method provides a framework for characterizing underlying or unidentified molecular changes or patterns.

## Data Availability

The spectral and medical data that support the findings of this study are available from the corresponding author upon reasonable request.

## Ethics Statement

All methods and experimental protocols were approved by the relevant institutional review boards (registration numbers 4453-12, 20-6830 and 17-6151, Ethics Commission, Faculty of Medicine, Ruhr-University Bochum). Furthermore, all enrolled patients provided informed consent that the samples may be used for retrospective analysis. All procedures were conducted in accordance with the approved guidelines and regulations for human experimental research.

## CRediT authorship contribution statement

**David Schuhmacher:** Methodology, Software, Validation, Formal analysis, Writing - Original Draft, Visualization. **Stephanie Schörner:** Software, Validation, Formal analysis, Investigation, Data Curation, Visualization. **Claus Kö pper:** Investigation, Writing - Review & Editing. **Frederik Großerueschkamp:** Writing - Review & Editing, Supervision. **Carlo Sternemann:** Data Curation. **Celine Lugnier:** Resources. **Anna-Lena Kraeft:** Resources. **Hendrik Jö tte:** Data Curation. **Andrea Tannapfel:** Resources, Data Curation, Writing - Review & Editing, Project administration, Funding acquisition. **Anke Reinacher-Schick:** Resources, Writing - Review & Editing, Project administration, Funding acquisition. **Klaus Gerwert:** Writing - Review & Editing, Supervision, Funding acquisition. **Axel Mosig:** Conceptualization, Methodology, Writing - Original Draft, Visualization, Supervision, Funding acquisition.

## Data availability

The spectral and medical data that support the findings of this study are available from the corresponding author AM upon reasonable request.

## Acknowledgements

The Center for Protein Diagnostics (PRODI) is funded by the Ministry of Culture and Science (MKW) of the State of North-Rhine Westphalia, Germany (grant number: 111.08.03.05-133974). Part of this research was conducted within the Slide2Mol project funded by the Computational Life Sciences program of the German Federal Ministry of Education and Research, grant number 031L0264.

## Supplement A: Implementation and Validation of Linear Relevance Propagation

−For comparison between the I-spaces induced by the CompSegNet on the one hand and linear relevance propagation (LRP) on the other hand, we applied LRP on a convolutional neural network trained on the same data from Kather et al. [2019a] as the CompSegNet. Following Kather et al. [2019b], we used a ResNet-18 He et al. [2016] architecture. For the sake of comparability with the CompSegNet, the ResNet-18 was not pretrained and initialized with random weights. The ResNet-18 was trained over 2,000 epochs on the same hardware as the CompSegNet. We implemented LRP by modifying an existing reference implementation Li, which supplements an early version of the implementation described in Lapuschkin et al. [2016] by relevance propagation through batch normalization layers. We also modified the relevance propagation through convolutional layers by utilizing tensorflow functionalities. Furthermore, we simplified the LRP in the ResNet topology by not taking into account the shortcut connections He et al. [2016] of the ResNet. The resulting relevance map of the input space of the ResNet-18 were handled in the spirit of Hägele et al. [2020]. First, the linear gradient approximations resulting from LRP require normalization. To this end, we determined minimum and maximum relevance across the 2,000 patches from the validation set and, following Hägele et al. [2020], normalized to a minimum relevance of − 1 and a maximum relevance of 1. The resulting three color channels of the input space were averaged to obtain a single relevance for each pixel position. We utilized the normalized LRP activation map as an intermediary I-space and determined a ROC curve over the same validation data set as the CompSegNet. Due to the relevance invariant underlying LRP Bach et al. [2015], the ROC curve of the ResNet-18 is equivalent to determining a ROC curve over the accumulated pixel relevances.

For computing Dice scores on the independent test set, LRP relevance maps were obtained by tiling the RoIs with overlapping patches. Subsequently, a relevance threshold was chosen to maximize the Dice score w.r.t. the manual ground truth segmentations, yielding the segmentations and Dice scores shown in Figure 7.

**Figure 7:**
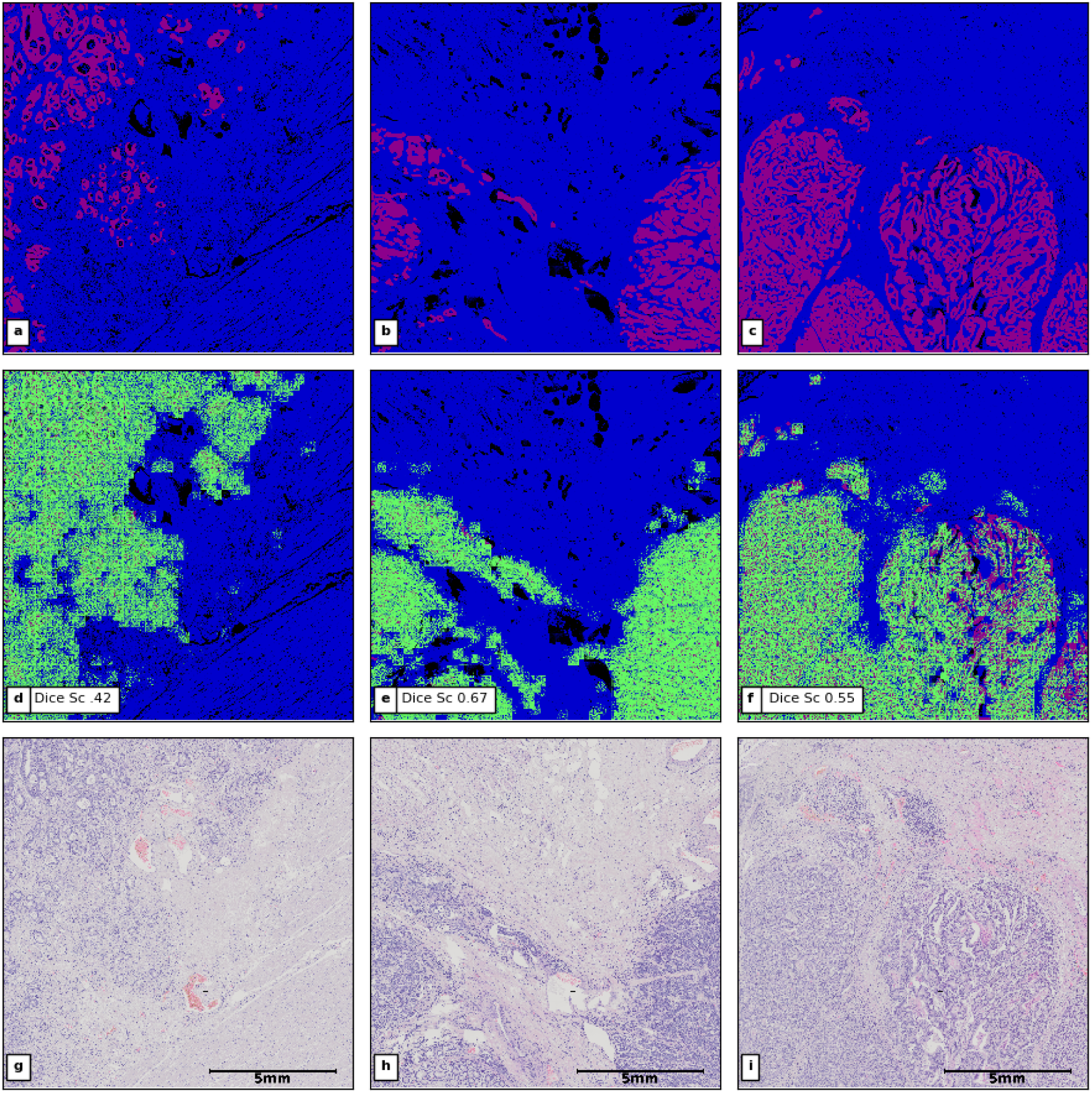
LRP-based segmentations of H&E stained images from the fully independent ColoPredict Plus 2.0 study test set. **a-c** Ground truth annotation of tumor and associated structures. Annotated tumor-associated regions are shown in purple, other tissue in blue. **d-f** Ground truth overlaid with binarized LRP relevance map (green). **g-i** H&E staining images underlying the annotation and the activation map.

Computation times for computing LRP relevance maps based on a trained ResNet-18 differ substantially from the computation times for computing activation maps based on a trained CompSegNet. Computing the relevance propagation to obtain an LRP relevance map in practice requires more than 1.8 seconds per patch. Obtaining the CompSegNet activation requires only the forward computation of the underlying U-Net, which in practice requires less than 0.05 seconds per patch. All practical computation times are observed on identical hardware (GPU node with four NVIDIA V100 cards and 128 GByte of GPU memory).

## References

Esther Abels, Liron Pantanowitz, Famke Aeffner, Mark D Zarella, Jeroen van der Laak, Marilyn M Bui, Venkata NP Vemuri, Anil V Parwani, Jeff Gibbs, Emmanuel Agosto-Arroyo, et al. Computational pathology definitions, best practices, and recommendations for regulatory guidance: a white paper from the digital pathology association. The Journal of Pathology, 249(3):286–294, 2019. doi: https://doi.org/10.1002/path.5331.

Mohamed Amgad, Habiba Elfandy, Hagar Hussein, Lamees A Atteya, Mai AT Elsebaie, Lamia S Abo El-nasr, Rokia A Sakr, Hazem SE Salem, Ahmed F Ismail, Anas M Saad, et al. Structured crowdsourcing enables convolutional segmentation of histology images. Bioinformatics, 35(18):3461–3467, 2019. doi: https://doi.org/10.1093/bioinformatics/btz083.

Sebastian Bach, Alexander Binder, Grégoire Montavon, Frederick Klauschen, Klaus-Robert Möller, and Wojciech Samek. On pixel-wise explanations for non-linear classifier decisions by layer-wise relevance propagation. PloS One, 10(7):e0130140, 2015. doi: https://doi.org/10.1371/journal.pone.0130140.

Harrison X Bai, Robin Wang, Zeng Xiong, Ben Hsieh, Ken Chang, Kasey Halsey, Thi My Linh Tran, Ji Whae Choi, Dong-Cui Wang, Lin-Bo Shi, et al. Artificial intelligence augmentation of radiologist performance in distinguishing COVID-19 from pneumonia of other origin at chest CT. Radiology, 296(3):E156–E165, 2020. doi: https://doi.org/10.1148/radiol.2020201491.

Wouter Bulten, Hans Pinckaers, Hester van Boven, Robert Vink, Thomas de Bel, Bram van Ginneken, Jeroen van der Laak, Christina Hulsbergen-van de Kaa, and Geert Litjens. Automated deep-learning system for Gleason grading of prostate cancer using biopsies: A diagnostic study. The Lancet Oncology, 21(2):233–241, 2020. doi: https://doi.org/10.1016/S1470-2045(19)30739-9.

Gabriele Campanella, Matthew G Hanna, Luke Geneslaw, Allen Miraflor, Vitor Werneck Krauss Silva, Klaus J Busam, Edi Brogi, Victor E Reuter, David S Klimstra, and Thomas J Fuchs. Clinical-grade computational pathology using weakly supervised deep learning on whole slide images. Nature Medicine, 25(8):1301–1309, 2019. doi: https://doi.org/10.1038/s41591-019-0508-1.

Yiming Ding, Jae Ho Sohn, Michael G Kawczynski, Hari Trivedi, Roy Harnish, Nathaniel W Jenkins, Dmytro Lituiev, Timothy P Copeland, Mariam S Aboian, Carina Mari Aparici, et al. A deep learning model to predict a diagnosis of Alzheimer disease by using 18F-FDG PET of the brain. Radiology, 290(2):456–464, 2019. doi: https://doi.org/10.1148/radiol.2018180958.

Juan Manuel Durán and Karin Rolanda Jongsma. Who is afraid of black box algorithms? On the epistemological and ethical basis of trust in medical AI. Journal of Medical Ethics, 47(5):329–335, 2021. doi: http://dx.doi.org/10.1136/medethics-2020-106820.

Frederik Großerueschkamp, Angela Kallenbach-Thieltges, Thomas Behrens, Thomas Bröning, Matthias Altmeier, Georgios Stamatis, Dirk Theegarten, and Klaus Gerwert. Marker-free automated histopathological annotation of lung tumour subtypes by FTIR imaging. Analyst, pages 2114–2120, 2015. doi: https://doi.org/10.1039/C4AN01978D.

Frederik Großerueschkamp, Thilo Bracht, Hanna C Diehl, Claus Kuepper, Maike Ahrens, Angela Kallenbach-Thieltges, Axel Mosig, Martin Eisenacher, Katrin Marcus, Thomas Behrens, et al. Spatial and molecular resolution of diffuse malignant mesothelioma heterogeneity by integrating label-free FTIR imaging, laser capture microdissection and proteomics. Scientific Reports, 7(1):1–12, 2017. doi: https://doi.org/10.1038/srep44829.

Riccardo Guidotti, Anna Monreale, Salvatore Ruggieri, Franco Turini, Fosca Giannotti, and Dino Pedreschi. A survey of methods for explaining black box models. ACM Computing Surveys, 51(5):1–42, 2018. doi: https://doi.org/10.1145/3236009.

Miriam Hägele, Philipp Seegerer, Sebastian Lapuschkin, Michael Bockmayr, Wojciech Samek, Frederick Klauschen, Klaus-Robert Möller, and Alexander Binder. Resolving challenges in deep learning-based analyses of histopathological images using explanation methods. Scientific Reports, 10(1):1–12, 2020. doi: https://doi.org/10.1038/s41598-020-62724-2.

Kaiming He, Xiangyu Zhang, Shaoqing Ren, and Jian Sun. Deep residual learning for image recognition. In Proceedings of the IEEE conference on computer vision and pattern recognition, pages 770–778, 2016.

Andreas Holzinger, Georg Langs, Helmut Denk, Kurt Zatloukal, and Heimo Möller. Causability and explainability of artificial intelligence in medicine. Wiley Interdisciplinary Reviews: Data Mining and Knowledge Discovery, 9(4): e1312, 2019. doi: https://doi.org/10.1002/widm.1312.

Ahmed Hosny, Chintan Parmar, Thibaud P Coroller, Patrick Grossmann, Roman Zeleznik, Avnish Kumar, Johan Bussink, Robert J Gillies, Raymond H Mak, and Hugo JWL Aerts. Deep learning for lung cancer prognostication: A retrospective multi-cohort radiomics study. PLoS Medicine, 15(11):e1002711, 2018. doi: https://doi.org/10.1371/journal.pmed.1002711.

Maximilian Ilse, Jakub Tomczak, and Max Welling. Attention-based deep multiple instance learning. In International conference on machine learning, pages 2127–2136. PMLR, 2018.

Angela Kallenbach-Thieltges, Frederik Großeröschkamp, Axel Mosig, Max Diem, Andrea Tannapfel, and Klaus Gerwert. Immunohistochemistry, histopathology and infrared spectral histopathology of colon cancer tissue sections. Journal of Biophotonics, 6(1):88–100, 2013. doi: https://doi.org/10.1002/jbio.201200132.

Angela Kallenbach-Thieltges, Frederik Großerueschkamp, Hendrik Jötte\, Claus Kuepper, Anke Reinacher-Schick, Andrea Tannapfel, and Klaus Gerwert. Label-free, automated classifcation of microsatellite status in colorectal cancer by infrared imaging. Scientific Reports, 10, 2020. doi: https://doi.org/10.1038/s41598-020-67052-z.

Jakob Nikolas Kather, Niels Halama, and Alexander Marx. 100,000 histological images of human colorectal cancer and healthy tissue. doi: http://dx.doi.org/10.5281/zenodo.1214456.

Jakob Nikolas Kather, Johannes Krisam, Pornpimol Charoentong, Tom Luedde, Esther Herpel, Cleo-Aron Weis, Timo Gaiser, Alexander Marx, Nektarios A Valous, Dyke Ferber, et al. Predicting survival from colorectal cancer histology slides using deep learning: A retrospective multicenter study. PLoS medicine, 16(1):e1002730, 2019a. doi: https://doi.org/10.1371/journal.pmed.1002730.

Jakob Nikolas Kather, Alexander T Pearson, Niels Halama, Dirk Jäger, Jeremias Krause, Sven H Loosen, Alexander Marx, Peter Boor, Frank Tacke, Ulf Peter Neumann, et al. Deep learning can predict microsatellite instability directly from histology in gastrointestinal cancer. Nature Medicine, 25(7):1054–1056, 2019b. doi: https://doi.org/10.1038/s41591-019-0462-y.

Christopher J Kelly, Alan Karthikesalingam, Mustafa Suleyman, Greg Corrado, and Dominic King. Key challenges for delivering clinical impact with artificial intelligence. BMC Medicine, 17(1):1–9, 2019. doi: https://doi.org/10.1186/s12916-019-1426-2.

Bruno Korbar, Andrea M Olofson, Allen P Miraflor, Catherine M Nicka, Matthew A Suriawinata, Lorenzo Torresani, Arief A Suriawinata, and Saeed Hassanpour. Looking under the hood: Deep neural network visualization to interpret whole-slide image analysis outcomes for colorectal polyps. In Proceedings of the IEEE Conference on Computer Vision and Pattern Recognition Workshops, pages 69–75, 2017.

Claus Kuepper, Frederik Großerueschkamp, Angela Kallenbach-Thieltges, Axel Mosig, Andrea Tannapfel, and Klaus Gerwert. Label-free classification of colon cancer grading using infrared spectral histopathology. Faraday Discussion, pages 105 – 118, 2016. doi: https://doi.org/10.1039/C5FD00157A.

Claus Kuepper, Angela Kallenbach-Thieltges, Hendrik Jötte, Andrea Tannapfel, Frederik Großerueschkamp, and Klaus Gerwert. Quantum cascade laser-based infrared microscopy for label-free and automated cancer classifcation in tissue sections. Scientific Reports, 8:1–10, 2018. doi: https://doi.org/10.1038/s41598-018-26098-w.

Sebastian Lapuschkin, Alexander Binder, Grégoire Montavon, Klaus-Robert Möller, and Wojciech Samek. The LRP toolbox for artificial neural networks. Journal of Machine Learning Research, 17(114):1–5, 2016. URL http://jmlr.org/papers/v17/15-618.html.

Sebastian Lapuschkin, Stephan Wäldchen, Alexander Binder, Grégoire Montavon, Wojciech Samek, and Klaus-Robert Möller. Unmasking Clever Hans predictors and assessing what machines really learn. Nature Communications, 10 (1):1–8, 2019. doi: https://doi.org/10.1038/s41467-019-08987-4.

Kedan Li. https://github.com/likedan/keraslrp.

Meng Li, Lin Wu, Arnold Wiliem, Kun Zhao, Teng Zhang, and Brian Lovell. Deep instance-level hard negative mining model for histopathology images. In International Conference on Medical Image Computing and ComputerAssisted Intervention, pages 514–522. Springer, 2019.

Zachary C Lipton. The mythos of model interpretability: In machine learning, the concept of interpretability is both important and slippery. Queue, 16(3):31–57, 2018.

Geert Litjens, Thijs Kooi, Babak Ehteshami Bejnordi, Arnaud Arindra Adiyoso Setio, Francesco Ciompi, Mohsen Ghafoorian, Jeroen Awm Van Der Laak, Bram Van Ginneken, and Clara I Sánchez. A survey on deep learning in medical image analysis. Medical Image Analysis, 42:60–88, 2017. doi: https://doi.org/10.1016/j.media.2017.07.005.

Ming Y Lu, Drew FK Williamson, Tiffany Y Chen, Richard J Chen, Matteo Barbieri, and Faisal Mahmood. Data-efficient and weakly supervised computational pathology on whole-slide images. Nature Biomedical Engineering, 5(6):555–570, 2021.

Marc Macenko, Marc Niethammer, James S Marron, David Borland, John T Woosley, Xiaojun Guan, Charles Schmitt, and Nancy E Thomas. A method for normalizing histology slides for quantitative analysis. In 2009 IEEE International Symposium on Biomedical Imaging: From Nano to Macro, pages 1107–1110. IEEE, 2009. doi: https://doi.org/10.1109/ISBI.2009.5193250.

Ryszard S Michalski. A theory and methodology of inductive learning. In Machine learning, pages 83–134. Elsevier, 1983. doi: https://doi.org/10.1016/B978-0-08-051054-5.50008-X.

Grégoire Montavon, Wojciech Samek, and Klaus-Robert Möller. Methods for interpreting and understanding deep neural networks. Digital Signal Processing, 73:1–15, 2018. doi: https://doi.org/10.1016/j.dsp.2017.10.011.

W James Murdoch, Chandan Singh, Karl Kumbier, Reza Abbasi-Asl, and Bin Yu. Definitions, methods, and applications in interpretable machine learning. Proceedings of the National Academy of Sciences, 116(44):22071–22080, 2019. doi: https://doi.org/10.1073/pnas.1900654116.

Arne P Raulf, Joshua Butke, Claus Köpper, Frederik Großerueschkamp, Klaus Gerwert, and Axel Mosig. Deep representation learning for domain adaptable classification of infrared spectral imaging data. Bioinformatics, 36(1): 287–294, 2020. doi: https://doi.org/10.1093/bioinformatics/btz505.

Olaf Ronneberger, Philipp Fischer, and Thomas Brox. U-net: Convolutional networks for biomedical image segmentation. In International Conference on Medical image computing and computer-assisted intervention, pages 234–241. Springer, 2015.

Ribana Roscher, Bastian Bohn, Marco F Duarte, and Jochen Garcke. Explainable machine learning for scientific insights and discoveries. IEEE Access, 8:42200–42216, 2020. doi: https://doi.org/10.1109/ACCESS.2020.2976199.

Cynthia Rudin. Stop explaining black box machine learning models for high stakes decisions and use interpretable models instead. Nature Machine Intelligence, 1(5):206–215, 2019. doi: https://doi.org/10.1038/s42256-019-0048-x.

Wojciech Samek, Grégoire Montavon, Sebastian Lapuschkin, Christopher J Anders, and Klaus-Robert Möller. Explaining deep neural networks and beyond: A review of methods and applications. Proceedings of the IEEE, 109 (3):247–278, 2021. doi: https://doi.org/10.1109/JPROC.2021.3060483.

David Schuhmacher, Klaus Gerwert, and Axel Mosig. A generic neural network approach to infer segmenting classifiers for disease-associated regions in medical images. medRxiv, 2020. doi: https://doi.org/10.1101/2020.02.27.20028845.

Leonie Selbach, Tobias Kowalski, Klaus Gerwert, Maike Buchin, and Axel Mosig. Shape decomposition algorithms for laser capture microdissection. Algorithms for Molecular Biology, 16(1):1–17, 2021. doi: https://doi.org/10.1186/s13015-021-00193-6.

Ramprasaath R Selvaraju, Michael Cogswell, Abhishek Das, Ramakrishna Vedantam, Devi Parikh, and Dhruv Batra. Grad-cam: Visual explanations from deep networks via gradient-based localization. In Proceedings of the IEEE international conference on computer vision, pages 618–626, 2017.

Karen Simonyan, Andrea Vedaldi, and Andrew Zisserman. Deep inside convolutional networks: Visualising image classification models and saliency maps. arXiv preprint 1312.6034, 2013.

Korsuk Sirinukunwattana, Josien PW Pluim, Hao Chen, Xiaojuan Qi, Pheng-Ann Heng, Yun Bo Guo, Li Yang Wang, Bogdan J Matuszewski, Elia Bruni, Urko Sanchez, et al. Gland segmentation in colon histology images: The GlaS challenge contest. Medical Image Analysis, 35:489–502, 2017. doi: https://doi.org/10.1016/j.media.2016.08.008.

Stanislau Trukhan, Valeria Tafintseva, Kristin Tøndel, Frederik Großerueschkamp, Axel Mosig, Vassili Kovalev, Klaus Gerwert, and Achim Kohler. Grayscale representation of infrared microscopy images by extended multiplicative signal correction for registration with histological images. Journal of Biophotonics, 13(8):e201960223, 2020. doi: https://doi.org/10.1002/jbio.201960223.

Jeroen van der Laak, Geert Litjens, and Francesco Ciompi. Deep learning in histopathology: The path to the clinic. Nature Medicine, 27(5):775–784, 2021. doi: https://doi.org/10.1038/s41591-021-01343-4.

Vladimir Vapnik. Estimation of dependences based on empirical data. pnSpringer, New York, 2006.

David H Wolpert. The lack of a priori distinctions between learning algorithms. Neural Computation, 8(7):1341–1390, 1996. doi: https://doi.org/10.1162/neco.1996.8.7.1341.

John R Zech, Marcus A Badgeley, Manway Liu, Anthony B Costa, Joseph J Titano, and Eric Karl Oermann. Variable generalization performance of a deep learning model to detect pneumonia in chest radiographs: A cross-sectional study. PLoS Medicine, 15(11):e1002683, 2018. doi: https://doi.org/10.1371/journal.pmed.1002683.

Bolei Zhou, Aditya Khosla, Agata Lapedriza, Aude Oliva, and Antonio Torralba. Learning deep features for discriminative localization. In Proceedings of the IEEE conference on computer vision and pattern recognition, pages 2921–2929, 2016.

